# Identification of Key Influencers for Secondary Distribution of HIV Self-Testing among Chinese MSM: A Machine Learning Approach

**DOI:** 10.1101/2021.04.19.21255584

**Authors:** Fengshi Jing, Yang Ye, Yi Zhou, Yuxin Ni, Xumeng Yan, Ying Lu, Jason J Ong, Joseph D Tucker, Dan Wu, Yuan Xiong, Chen Xu, Xi He, Shanzi Huang, Xiaofeng Li, Hongbo Jiang, Cheng Wang, Wencan Dai, Liqun Huang, Wenhua Mei, Weibin Cheng, Qingpeng Zhang, Weiming Tang

## Abstract

**Background:** HIV self-testing (HIVST) has been rapidly scaled up and additional strategies further expand testing uptake. Secondary distribution has people (indexes) apply for multiple kits and pass these kits to people (alters) in their social networks. However, identifying key influencers is difficult. This study aimed to develop an innovative ensemble machine learning approach to identify key influencers among Chinese men who have sex with men (MSM) for HIVST secondary distribution.

**Method:** We defined three types of key influencers: 1) key distributors who can distribute more kits; 2) key promoters who can contribute to finding first-time testing alters; 3) key detectors who can help to find positive alters. Four machine learning models (logistic regression, support vector machine, decision tree, random forest) were trained to identify key influencers. An ensemble learning algorithm was adopted to combine these four models. Simulation experiments were run to validate our approach.

**Results:** 309 indexes distributed kits to 269 alters. Our approach outperformed human identification (self-reported scales cut-off), exceeding by an average accuracy of 11·0%, could distribute 18·2% (95%CI: 9·9%-26·5%) more kits, find 13·6% (95%CI: 1·9%-25·3%) more first-time testing alters and 12·0% (95%CI: -14·7%-38·7%) more positive-testing alters. Our approach could also increase simulated intervention efficiency by 17·7% (95%CI: -3·5%-38·8%) than human identification.

**Conclusion:** We built machine learning models to identify key influencers among Chinese MSM who were more likely to engage in HIVST secondary distribution.

**Key Findings (can also be found in Figure.2-Infographic):** Our proposed ensemble machine learning approach outperformed human identification (self-reported scales cut-off) in accuracy & *F_1_* by classification metrics and in intervention efficiency by simulation experiments. Our model could also distribute more kits, find more first-time/positive-testing alters than human identification.

## 1 Introduction

Men who have sex with men (MSM) have a higher burden of HIV (1). In China, HIV prevalence among MSM is 6·3% in 2019 (2). However, over 40% of Chinese MSM have never been tested (3) and over 30% of MSM living with HIV do not know their serostatus (4). More efficient case-finding for undiagnosed people living with HIV and starting treatment is essential for HIV control (5). To increase HIV testing coverage, HIV self-testing (HIVST) has been recommended by the World Health Organization (WHO) (6), which has high acceptability among MSM (7).

Secondary distribution is one of the novel ways to increase the use of HIVST (8). In this service delivery model, individuals (defined as indexes) apply for multiple HIVST kits and then distribute these HIVST kits to people in their social network, such as sexual partners and gay friends (defined as alters) (9,10). Such a strategy could significantly improve HIV testing coverage by reaching people who have limited access to HIV testing and potentially detect more people with undiagnosed HIV (10). To further expand the use of this strategy and enhance the efficiency of distribution, it could be useful to identify influential indexes who are more likely to distribute kits to more alters (e.g., ≥2 alters), or to people living with HIV (PLWH) who are undiagnosed, or to first-time testers.

However, existing methods for identifying MSM key influencers are limited in the following two respects. First, some studies selected key influential people based on human intuition and then trained them as opinion leaders (11,12). This selection process of key influencers lacks a scientific basis and is not reliable or generalizable (13). Second, other studies utilized self-reported leadership scales, such as among drug users (14). This method is more scientific because of self-reported leadership scales, but it is still relatively subjective. Even if all self-reported leadership items were reliable and valid, these identified leaders in the community might not be key influencers for HIVST secondary distribution. Artificial intelligence (AI) including machine learning (ML) approaches is a promising method to identify key influencers (15,16). In the area of HIV intervention, machine learning models also performed well in different kinds of key population classification tasks, like identifying people with relatively higher risk of HIV (17) and identifying suitable candidates for pre-exposure prophylaxis (PrEP) (18). Thus, machine learning approaches have potential to be used for identifying key influencers for secondary distribution of HIVST kits.

Using data collected from previous studies (19), we proposed a novel ensemble machine learning approach (Figure 1) to identify key influencers for secondary distribution of HIVST kits where indexes applied for testing kits for distribution while alters were those who received these kits. Specifically, our machine learning models were trained to obey such three rules in order to identify key influencers: key-distribution influencers (i.e., key distributors) who are more likely to distribute kits to as many alters as possible (e.g., no fewer than two kits in ten months); key-promotion influencers (i.e., key promoters) who contribute to promoting first-time testing among alters; key-detection influencers (i.e., key detectors) who distribute kits to alters who are undiagnosed people living with HIV (PLWH).

**Figure 1.**
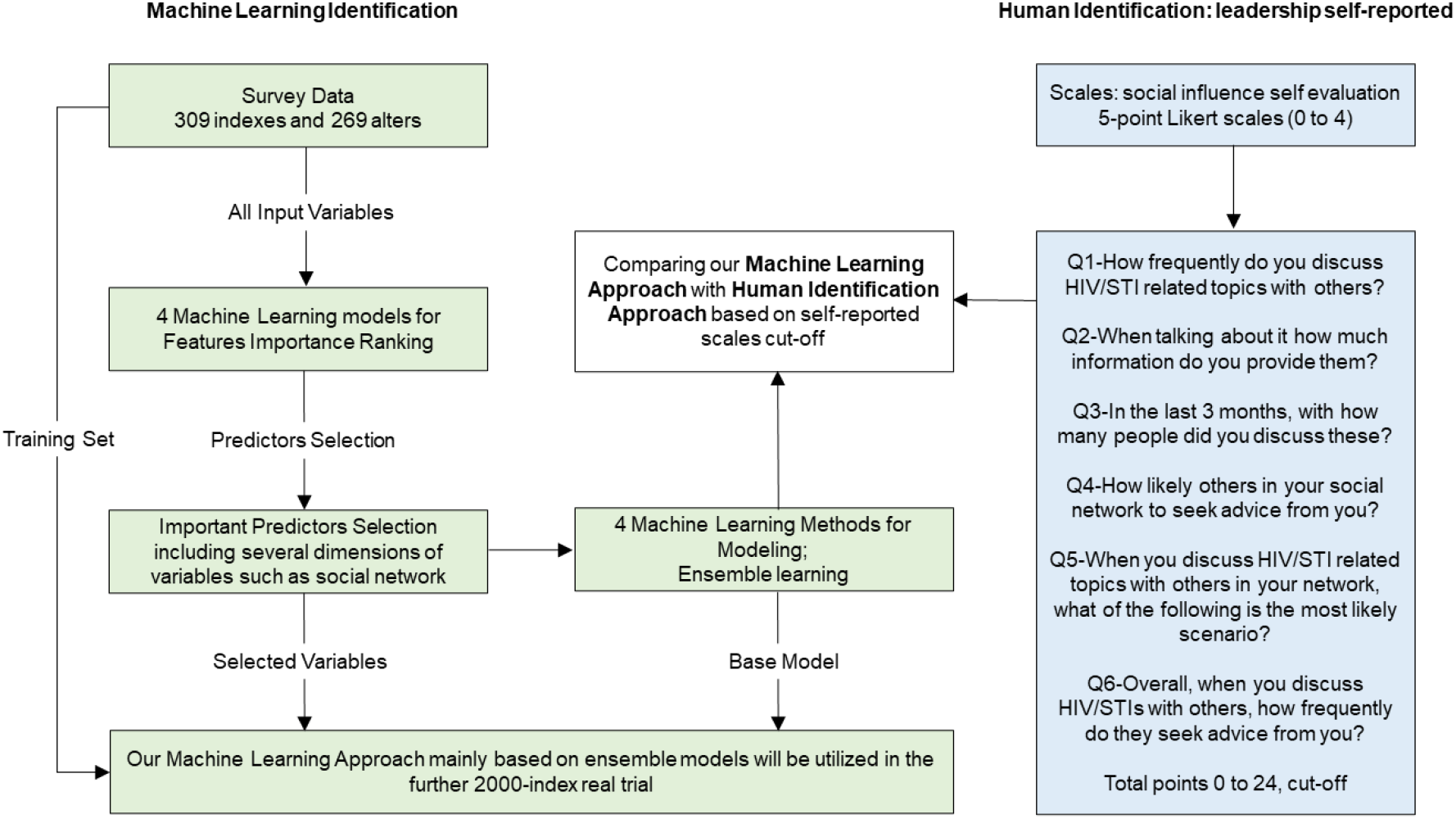
Framework of our study.

**Figure 2.**
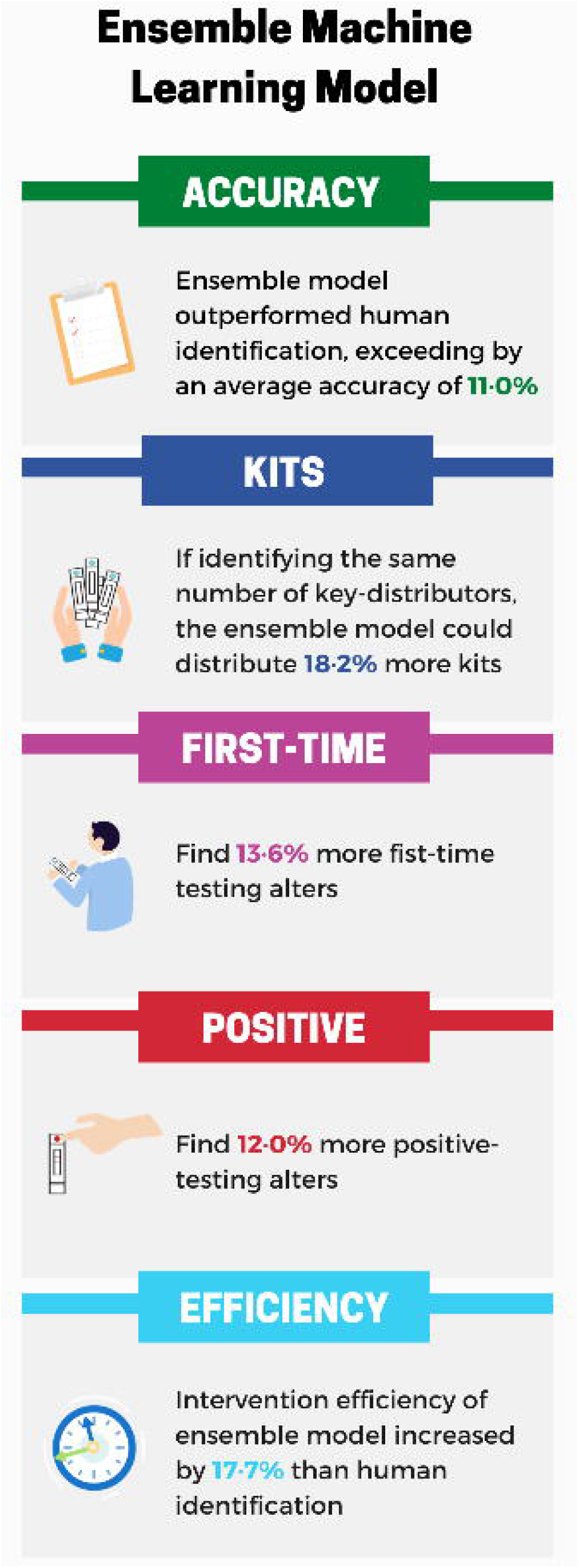
Infographic.

## 2 Methods

### 2.1 Data Processing

The dataset was derived from a three-arm randomized control trial of HIVST secondary distribution in Zhuhai, China (19). In this trail, 309 MSM were recruited as indexes and randomly assigned to the control group (standard secondary distribution arm), the intervention I group (secondary distribution with monetary incentives arm), or the intervention II group (secondary distribution with monetary incentives plus peer referral). Of 309 indexes, 60 were key distributors who passed the kits to at least two alters. Additionally, there were 73 key promoters leading to 103 alters who were first -time testers, and 23 key detectors leading to 25 alters who were undiagnosed PLWH, as defined above.

### 2.2 Machine Learning Modeling

We formulated the key influencers identification as a binary classification problem and four machine learning models were constructed, including logistic regression, support vector machine, decision tree and random forest (20,21). Each model has its pros and cons (see Supplement A) in performing the classification task, and thus we comprehensively combined the predictions of all four models to mitigate overfitting and model biases by adopting an ensemble learning approach (22), which could synthesize the strengths from each machine learning model.

As for how to evaluate machine learning models for such classification tasks, we used four metrics for performance evaluation: accuracy, precision, recall and F1-score (See Table S1). Accuracy is defined as a ratio of correctly predicted observations to the total observations. The F1-score takes both precision and recall into consideration.

We employed five-fold cross-validation (23) to ensure the robustness of the models and compared the average values of each metric. Specifically, we randomly sampled 80% of data for training and 20% of data for testing. Experiments for each metric were repeated five times as every time one fold (20% of data) would be the testing set and the remaining four folds (80% of data) would be trained for model construction and parameters learning. The final average values of each metric are the average performance of the five folds’ testing set.

### 2.3 Predictors Selection

First, we incorporated original predictors (i.e., input variables from survey) into our classification models using the aforementioned four machine learning models. Then we obtained a predictor ranking list of each machine learning model ordered by the importance of every variable (see Supplement B). The four machine learning models voted for the final selected predictors, which ranked top in all four importance ranking lists.

### 2.4 Identification System

After determining the top predictors by importance ranking, we ran the same four machine learning models based on these selected variables to check and obtain the classification performance. Then we used ensemble learning to combine the findings from all machine learning models. The whole process, including machine learning modeling, predictors selection, and modeling based on selected important variables, ensemble learning, represents a novel intelligent identification system (illustrated in Figure 1), which can be adopted in HIVST secondary distribution implementation program in the future to identify key influencers among MSM.

We ran all machine learning experiments in Python 3.7 and the code is available through this link: https://sites.google.com/view/jingfengshi/home/blog/code for sharing.

### 2.5 Human Identification Approach

To compare with our machine learning results, we used two self-reported scales as the human identification approach. According to existing literature, self-evaluated leadership scales are commonly used to identify key influencers (11,12,14). These self-reported leadership scales asked indexes to evaluate the likelihood of six social influence-related scenarios on a scale of 0-4; the total points range from 0 to 24. Based on the previous literature, indexes (around 20% out of total 309 indexes) who distributed at least two kits were defined as key distributors on the ground truth. Hence, we should also cut off the top 60 indexes (also around 20%) by the ranking order of our 6-question self-reported scales (Figure 1) as the human identified key influencers. However, 49 indexes got at least 11 points in the self-reported scales while 81 indexes got at least 10 points. In other words, since the 49th to the 80th indexes got the same points in the scales, we were unable to determine exactly whom of these indexes belonged to the top 60. As a result, we regarded these two scale cut-offs as human identification baselines together, recorded as cut-off A and cut -off B respectively.

### 2.6 Simulation

Finally, we further conducted a simulation model to mimic the secondary distribution process on the MSM social network and compare the intervention efficiency of key influencers identified by the machine learning models and the conventionally human identification approaches. Here, the intervention efficiency is defined as the number of individuals who have tested themselves at the end of the simulation. Specifically, simulation technologies (24,25) on HIV-related networks (26,27) can also model distribution network characteristics.

We simulated the secondary distribution process on each test set of the five-fold cross-validation. Given indexes of each test set, we can construct a network containing both indexes and alters who received self-testing kits from these indexes. There would be an edge between an index and an alter if the alter received a kit with the corresponding confirmation code of the index. Self-testing kits would be distributed through edges on the network. As illustrated in Table 3, we observe a higher distribution efficiency for machine learning models than that for conventionally human identification approaches. More technical det ails of this simulation are presented in the Supplemental Materials.

## 3 Results

### 3.1 Modeling Results

We compared the machine learning classification performance and the ensemble learning one, with the human identification approach based on leadership self-evaluated scales in terms of the two nearest cut-offs. In our survey data, 60 indexes (19·4%, around 20%) who distributed at least two kits were key distributors on the ground truth. In addition, these key distributors reached more than 70% of alters in total. Additionally, there were 73 key promoters who helped us to promote 103 alters for first -time testing and 23 key detectors who helped us to detect 25 positive alters.

Table 1 illustrates that machine learning classification results significantly outperformed human identification cut-off no matter which type (i.e., key distributors, key promoters and key detectors) was adopted to define the key influencers. The model using ensemble learning, also outperformed the human identification approach and nearly achieved the highest value among all models in terms of the performance metrics. Specifically, for three classification training rules (i.e., three types of key influencers), the classification performance of ensemble learning obtained an accuracy of 90%, 93% and 82% respectively, all exceeding human identification (self-reported scales cut-off). Therefore, the ensemble learning approach combining four machine learning models was able to better capture key influencers, compared with the other approaches studied, exceeding by an average accuracy of 11·0% than human identification.

**Table 1.** Machine learning classification results using five-fold cross-validation.

| Metrics | Key Distributors<br>Number of Alters $\geq 2$ | | Key Detectors<br>Positive Alters $\geq 1$ | | Key Promoters<br>New-Tester Alters $\geq 1$ | |
| --- | --- | --- | --- | --- | --- | --- |
| | Accuracy | $F_1$-score | Accuracy | $F_1$-score | Accuracy | $F_1$-score |
| ***LR*** | 0.89 | 0.93 | 0.92 | 0.95 | 0.83 | 0.89 |
| ***SVM*** | 0.90 | 0.94 | 0.93 | 0.96 | 0.82 | 0.89 |
| ***DT*** | 0.91 | 0.94 | 0.91 | 0.95 | 0.80 | 0.88 |
| ***RF*** | 0.88 | 0.93 | 0.93 | 0.96 | 0.78 | 0.87 |
| ***Ensemble*** | 0.90 | 0.94 | 0.93 | 0.96 | 0.82 | 0.89 |
| ***Cut-off A*** | 0.72 | 0.82 | 0.85 | 0.88 | 0.68 | 0.78 |
| ***Cut-off B*** | 0.79 | 0.87 | 0.88 | 0.91 | 0.73 | 0.83 |

LR represents for Logistic Regression; SVM represents for Support Vector Machine; DT represents for Decision Tree; RF represents for Random Forest; Ensemble represents for Ensemble Learning; Cut-off A means self-reported scales lower cut-off while Cut-off B means upper cut-off of self-reported scales. Five-fold cross-validation technical details can be found in Supplement.

Table 2 compares the number of distributed kits from key influencers identified using the ensemble learning model and the human identification approach. Both the ensemble learning and human identification approach classified 49 key influencers each but those identified using the ensemble learning approach distributed 146 kits, equating to 54% of alters (i.e., 146 alters out of 269). In contrast, the 49 key influencers identified using the human identification approach only distributed 97 kits. In addition, the same 49 key influencers identified by ensemble learning found 3 more PLWH and 14 more first -time testers than key influencers identified by human identification. In summary, our new approach could distribute 18·2% (95% CI: 9·9%-26·5%) more kits, find 13·6% (95% CI: 1·9%-25·3%) more first-time testing alters, and 12·0% (95% CI: -14·7%-38·7%) more undiagnosed PLWH.

**Table 2.** Key-distributing influencers ensemble learning identification results and its comparison.

| Number of | Successfully distributed kits | Positive alters (i.e., PLWH) | First-time testing alters |
| --- | --- | --- | --- |
| *ML Identification* | 146 | 11 | 33 |
| *Self-reported Scales* | 97 | 8 | 19 |
| *Total (Original)* | 269 | 25 | 103 |
| *Increased percentage* | 18·2% | 12·0% | 13·6% |

The above results are based on ensemble learning model for identifying key -distributing influencers (i.e., key distributors), and such model happened to classify 49 key influencers (in five-fold testing sets), sharing the same number with one certain scales cut-off identification. Therefore, our comparisons are rational. Note that such percentages and confidence intervals are calculated based on the total number (e.g., there're total 269 distributed kits, hence the increased percentage of successfully distributed kits is calculated through the equation $(146-97)/269$ instead of another equation $(146-97)/146$ ).

### 3.2 Simulation Results

We simulated the secondary distribution process on each test set of five-fold cross-validation. The simulation results (Table 3) demonstrated that our ensemble machine learning approach could always obtain a higher intervention efficiency in each fold than the conventional human identification approach. Specifically, the average intervention efficiency of ensemble machine learning model increased by 17·7% (95% CI: -3·5%-38·8%) than self-reported scales cut-off method, which indicated the higher intervention efficiency of our novel method to identify key influencers.

**Table 3.** Simulation results of distribution efficiency after key influencers being identified.

| Efficiency | Fold 1 | Fold 2 | Fold 3 | Fold 4 | Fold 5 | Average |
| --- | --- | --- | --- | --- | --- | --- |
| ***ML*** | 72·7% | 68·5% | 65·0% | 75·0% | 79·6% | 72·2% |
| ***Human*** | 64·6% | 58·0% | 51·3% | 36·8% | 61·9% | 54·5% |

ML means ensemble machine learning identification results, while Human means self-reported scales cut-off results.

## 4 Discussion

Identifying key influencers for secondary distribution of HIVST among Chinese MSM is important. Identification of key influencers who may be more active in HIVST secondary distribution can potentially expand testing coverage, reach more naïve testers, and help identify undiagnosed PLWH. We found that using a machine learning approach (specifically ensemble learning) was superior to using human identification of key influencers.

We found that all four machine learning models achieved higher performance than the human identification approach. Our results are consistent with other studies that machine learning models have performed better in other HIV-related identification tasks like identifying high-risk HIV populations (17), identifying HIV-related social media data (28), and identifying people eligible for PrEP (18,29). This adds to the evidence showing that machine learning can be used to identify key influencers within networks more efficiently than other methods.

In addition, we found that key influencers identified by our ensemble learning approach could distribute 18·2% more kits, find 13·6% more first-time testing alters and 12·0% more undiagnosed PLWH than the conventional human identification approach. As for why our ensemble learning approach outperformed the human identification approach (14), we believe this may be because the machine learning algorithms included variables on two key drivers for identification: index men’s HIV testing and kits application. Self-reported leadership scales are not designed to specifically consider such important predictors. Our data suggest that ML could improve on social evaluation scales to identify key influencers. Using a machine learning approach could enhance the public health impact of secondary distribution.

Our method provides one potential means to prioritize indexes identified as key influencers in HIVST secondary distribution using machine learning. Our novel ensemble machine learning approach for key influencers identification of HIVST secondary distribution can accurately and quickly classify which indexes are important for distributing more kits, promoting testing among more naive testers or detecting more undiagnosed PLWH. This is particularly important for low-and middle-income countries (LMIC) where resources for HIV testing services may be relatively limited.

Our study also has several limitations. First, our study was a retrospective modeling research, and a prospective trial to compare machine learning and conventional methods is needed. Second, due to the survey content, we only compared our machine learning approach with one kind of human identification, that is, self-reported leadership scales cut-off. Future studies should compare the machine learning approach with the other methods for human identification.

In conclusion, we found that machine learning using ensemble learning achieved the highest accuracy in identifying key influencers who are more effective at secondary distribution of HIVST in China. Thus, for future research plans, we will adopt this approach in our future programs of 2000-index HIVST secondary distribution, and a randomized controlled trial (machine learning identification v.s. scales-based human identification). Our ensemble learning approach can also be generalized to identify key influencers for other HIV prevention and treatment programs.

## Data Availability

Training set data for machine learning modeling will be made available to others after obtaining the relevant data sharing agreement and finishing the future quasi-experiment.

## Acknowledgement

We thank all study participants, Zhuhai Xutong CBO staff, Katherine T Li, Haidong Lu, and Jie Fan for their invaluable support.

## Contributors

FJ, YY and YZ drafted the manuscript together, while FJ contributed to machine learning modeling part and YY contributed to simulation experiment part. YZ, JJO, JDT, DW, WC, QZ, and WT provided oversight and made insightful contributions to the study conception. YN, YL, CX, XH, XL, and SH collected and cleaned original survey data. YN, XY, JJO, JDT, DW, YX, HJ, CW, WD, LH, WM, WC, QZ, and WT critically revised the paper. All authors reviewed and authorized the final manuscript.

