## Supplements for "Identification of Key Influencers for Secondary Distribution of HIV Self-Testing among Chinese MSM: A Machine Learning Approach"

**Supplementary Materials**

Supplementary Technical Details (Supplement A to F)

Supplementary References (SR1-SR5)

Supplementary Tables (Table S1 & S2)

### Supplementary Technical Details (Supplement A to F)

#### Supplement A: Four machine learning models explanation

The logistic regression (LR) model is more suitable to survey data where categorical variables are quite common (SR1). The support vector machine (SVM) model is also suitable for classification tasks as another machine learning method (SR2). The decision tree (DT) model is just like a human thinking process to make a decision (SR3). Random forest (RF) model can be simply understood as a Bootstrap aggregating Classification And Regression Trees (CART) algorithm based on decision trees (SR4).

#### Supplement B: Features importance ranking algorithms

The logistic regression model provided us with the coefficient of each predictor while the other three models (support vector machine, decision tree, and random forest) provides the importance level. Specifically, in logistic regression, we rank the importance of each characteristic according to the absolute value of the standardized coefficient. For support vector machine model, we adopted the Recursive Feature Elimination (RFE) (SR5) algorithm to generate a weighted vector when training and then in each time of iteration, we eliminated a least important feature through the above weighted vector. For decision tree and random forest, GINI index in CART algorithm (SR4) determined the importance of every variable.

#### Supplement C: Selected predictors results analysis

We finished predictors selection (results are shown in Table S2). After we obtained four lists of importance ranking from 4 machine learning models, we calculated the occupancy percentage of each predictor's interpretability in each model. We ranked the overall predictors according to the sum of occupancy percentage from 4 machine learning models. Among those selected variables, HIV testing and kits application information of each index is the most important because our key influencers identification is in respect of the secondary distribution of HIVST kits, such as how many kits does the index would like to request, does the index plan to give the kit to his partners, and so on. Additional scales only capture self-reported leadership, which is indeed an indispensable aspect, but the influencers what we aim to identify is who have greatest influence in the secondary distribution of HIVST kits. Therefore, the information of HIV testing and kits application playing a key role makes sense. It discloses that in connection with different objectives for key influencers identification, we are not supposed to use the same self-reported scales which may not be completely consistent with characteristics what we really want to identify.

### Supplement D: Predictors selection discussion

We successfully identified 12 predictors based on the second survey, which can be further implemented in future studies. Variables about social network and whether belonging to control/intervention groups were selected as important predictors. This is in accord with common sense since monetary incentives ought to drive people to distribute more kits and social network connection decides whether the index has enough partners for possible distribution behaviors. In summary, these two dimensions of variables were previously overlooked in the first survey and would make a difference in identification for similar trials in the future.

### Supplement E: Five-fold cross-validation

We employed five-fold cross-validation to ensure the robustness of the models and compared the average values of each metric. Specifically, we randomly sampled 80% of data for training and 20% of data for testing. Those 80% data are equal to four folds and 20% data are equal to one fold. Five folds are mutually exclusive and exhaustive. Experiments for each metric were repeated five times as every time one fold would be testing set and the remaining four folds would be trained for model construction and parameters learning. The final average values of each metric are the five-time average performance of the testing set.

### Supplement F: Simulation details

We describe the details of the secondary distribution process. In the beginning, all nodes have not tested themselves. Key influencers are provided with k =4 kits. Nodes who have kits and have not tested themselves will use one kit for self-testing. Then they will distribute the remaining kits to their neighbors on the social network. The number of kits that they want to distribute to others obeys a Poisson distribution. The mean number of kits that they want to distribute is equal to the number of kits they have. For example, a node has k kits after self-testing, then the number of kits that it wants to distribute to neighbors obeying a Poisson distribution. Assuming that each node can know if neighbors have tested themselves through communication. Nodes will first distribute kits to those who have not tested themselves, then distribute the remaining kits to neighbors equally. If kits are not available to distribute to all neighbors, the node will distribute kits to some randomly selected neighbors. Nodes who have kits and have not tested themselves will use one kit for self-testing. According to this scheme, we can get the distribution efficiency of machine learning identification and human identification as follows. Results are obtained by averaging 1000 simulations.

(SR5) Bedo, J., Sanderson, C., & Kowalczyk, A. (2006, December). An efficient alternative to svm based recursive feature elimination with applications in natural language processing and bioinformatics.

In Australasian Joint Conference on Artificial Intelligence (pp. 170-180). Springer, Berlin, Heidelberg.

**Table S1. Metrics definition, explanation, and formulas in classification performance evaluation**

| Definition | **Meaning and Formula** |
|---|---|
| ***TP*** | True Positive |
| ***FP*** | False Positive |
| ***FN*** | False Negative |
| ***TN*** | True Negative |
| ***Accuracy*** | (TP+TN)/(TP+FP+FN+TN) |
| ***Precision*** | TP/(TP+FP) |
| ***Recall (i.e., Sensitivity)*** | TP/(TP+FN) |
| ***$F_1$-score*** | 2×Precision×Recall / (Precision + Recall) |

Actually, from mathematical perspective, F1-score is the harmonic mean of Precision and Recall. That is, we can obtain F1-score by 2/F1 = 1/Precision + 1/Recall. Accuracy and F1-score are most widely used in classification tasks as two comprehensive metrics.

**Table S2. Predictors Selection**

| Selected Predictors | **Variable Dimension** | **Variable Types** |
|---|---|---|
| How many self-test kits do you want to request this time? | HIV testing and kits application | Numeric variable |
| Do you plan to give the kit to others? | HIV testing and kits application | Categorical variable |
| Do you want to get the promotion link of the kit? | HIV testing and kits application | Categorical variable |
| How did you test for HIV the previous time? | HIV testing and kits application | Categorical variable |
| Arm (which groups) | Controlled or intervention group | Categorical variable |
| How frequently do you have sex with her/him? | Social network | Categorical variable |
| What is her/his sexual self-identification (gender) | Social network | Categorical variable |
| In the last 3 months, how many stable male partners did you have? | MSM behavior | Numeric variable |
| In the last 3 months, how many casual male partners did you have? | MSM behavior | Numeric variable |
| In the last 3 months, how frequently did you use the new drugs? | MSM behavior | Categorical variable |
| Overall, how frequently do you discuss HIV/STI related topics with others? | Self-reported leadership scales | Scales variable |
| When you discuss HIV/STI related topics with others in your network, what of the following is the most likely scenario? | Self-reported leadership scales | Scales variable |
